# Host genomics of the HIV-1 reservoir size and its decay rate during suppressive antiretroviral treatment

**DOI:** 10.1101/19013763

**Authors:** Christian W. Thorball, Alessandro Borghesi, Nadine Bachmann, Chantal von Siebenthal, Valentina Vongrad, Teja Turk, Kathrin Neumann, Niko Beerenwinkel, Jasmina Bogojeska, Volker Roth, Yik Lim Kok, Sonali Parbhoo, Mario Wieser, Jürg Böni, Matthieu Perreau, Thomas Klimkait, Sabine Yerly, Manuel Battegay, Andri Rauch, Patrick Schmid, Enos Bernasconi, Matthias Cavassini, Roger D. Kouyos, Huldrych F. Günthard, Karin J. Metzner, Jacques Fellay, Swiss HIV Cohort Study

## Abstract

**Introduction:** A major hurdle to HIV-1 eradication is the establishment of a latent viral reservoir early after primary infection. Several factors are known to influence the HIV-1 reservoir size and decay rate on suppressive antiretroviral treatment (ART), but little is known about the role of human genetic variation.

**Methods:** We measured the reservoir size at three time points over a median of 5.4 years, and searched for associations between human genetic variation and two phenotypic readouts: the reservoir size at the first time point and its decay rate over the study period. We assessed the contribution of common genetic variants using genome-wide genotyping data from 797 patients with European ancestry enrolled in the Swiss HIV Cohort Study and searched for a potential impact of rare variants and exonic copy number variants using exome sequencing data generated in a subset of 194 study participants.

**Results:** Genome- and exome-wide analyses did not reveal any significant association with the size of the HIV-1 reservoir or its decay rate on suppressive ART.

**Conclusions:** Our results point to a limited influence of human genetics on the size of the HIV-1 reservoir and its long-term dynamics in successfully treated individuals.

## INTRODUCTION

Combination antiretroviral treatment (ART) has turned the previously lethal infection by human immunodeficiency virus type 1 (HIV-1) into a chronic disease. Despite this significant achievement, HIV-1 as retrovirus, self-integrating its genome into the host chromosome, persists indefinitely in infected individuals during treatment [1–4], and life-long ART is required to control the infection.

A major hurdle to HIV-1 eradication is the establishment, already during primary infection, of a latent viral reservoir of HIV-1 DNA persisting as provirus in resting memory CD4^+^ T cells [1,2,5–8]. At the molecular level, chromatin remodeling, epigenetic modifications, transcriptional interference, and availability of transcription factors have been considered as possible mechanisms contributing to HIV-1 latency [9]. The viral reservoir is measurable through different methods, including viral outgrowth assay and intracellular HIV-1 DNA quantification [10,11]. Currently, there is no consensus on the best HIV-1 reservoir biomarker. Total cell-associated HIV-1 DNA, easy to measure in different cell and tissue samples and applicable to large populations, has been shown to be a good proxy for the reservoir size [12]. Indeed, while HIV-1 DNA measurement is able to detect both integrated and nonintegrated viral genomes coding for intact or defective viruses [13], total HIV-1 DNA levels have been shown to correlate with viral outgrowth [14], and to predict the time to viral rebound at treatment interruption [15]. Moreover, the substantial loss of nonintegrated HIV-1 DNA genomes following ART initiation suggests that total HIV-1 DNA after prolonged suppression is largely accounted for by integrated viral genomes [16].

After an initial rapid decay following ART initiation, changes of the viral reservoir size over time display wide inter-individual variability. By limiting dilution culture assay, the half-life of the viral reservoir was first estimated to be 44 months (95% confidence interval 27.4-114.5) in individuals with undetectable viremia [4]. A more recent study showed a slow decline of total HIV-1 DNA with a half-life of 13 years after the first four years of suppressive ART [17]. Generally, different studies show a broad variability of the average decay rate, from 2.5 months to no measurable decay [18–26]. One study even reported an increase in the viral reservoir size in as much as 31% of patients in the 4-7 years following ART initiation [27], and recent data from our group confirm this observation, reporting an increase in the reservoir size in 26.8% of individuals in the 1.5-5.5 years after ART initiation [28].

Several factors are known to influence the decay rate of the viral reservoir: initiation of ART during acute HIV-1 infection substantially accelerates the decay rate, while viral blips and low-level viremia during ART slow it down, as shown in previous studies [22] and in recent data from our cohort [28]. Conversely, treatment intensification, i.e. treating with additional drugs, does not appear to influence the decay rate, suggesting that residual replication is not the main driver of the viral reservoir [29] or that it may happen in sanctuary sites.

Human genetic variants have been shown to influence the outcome of various infections, including HIV. Previous genome-wide association studies (GWAS) addressed the role of common genetic polymorphisms in several HIV-related phenotypes, including plasma viral load (HIV-1 RNA) at set point, exceptional capacity to control viral replication, pace of CD4+ T lymphocyte decline, time to clinical AIDS, rapid progressor status or long-term non-progressor status (LTNP) [30–37], and, in one single study, the amount of intracellular HIV-1 DNA, measured at a single time point during the chronic phase of infection [38]. Rare genetic variants that are detectable through DNA sequencing technologies have been investigated far less. However, a large exome sequencing study did not reveal any convincing association of such variants with the natural history of HIV disease [39].

To date, no studies have addressed the role of human genetic variation in determining the initial viral reservoir size and the reservoir decay rate over time. In the current study, we searched for host genetic factors associated with the HIV-1 reservoir size and its long-term dynamics in a cohort of 797 HIV-1 positive individuals on suppressive ART for at least five years.

## METHODS

### Ethics statement

Participants in the Swiss HIV Cohort Study (SHCS) consented to the cohort study and genetic analyses, as approved by the corresponding local Ethics Committees.

### Study participants

The SHCS is an ongoing, nation-wide cohort study of HIV-positive individuals, including more than 70% of all persons living with HIV in Switzerland. Clinical and laboratory information has been prospectively recorded at follow-up visits every 3-6 months since 1988 [40]. The general enrolment criteria have been described previously [28]. Additionally, availability of genome-wide genotyping data from previous studies or of a DNA sample for genotyping was required for inclusion in this study (Figure 1).

**Figure 1.**
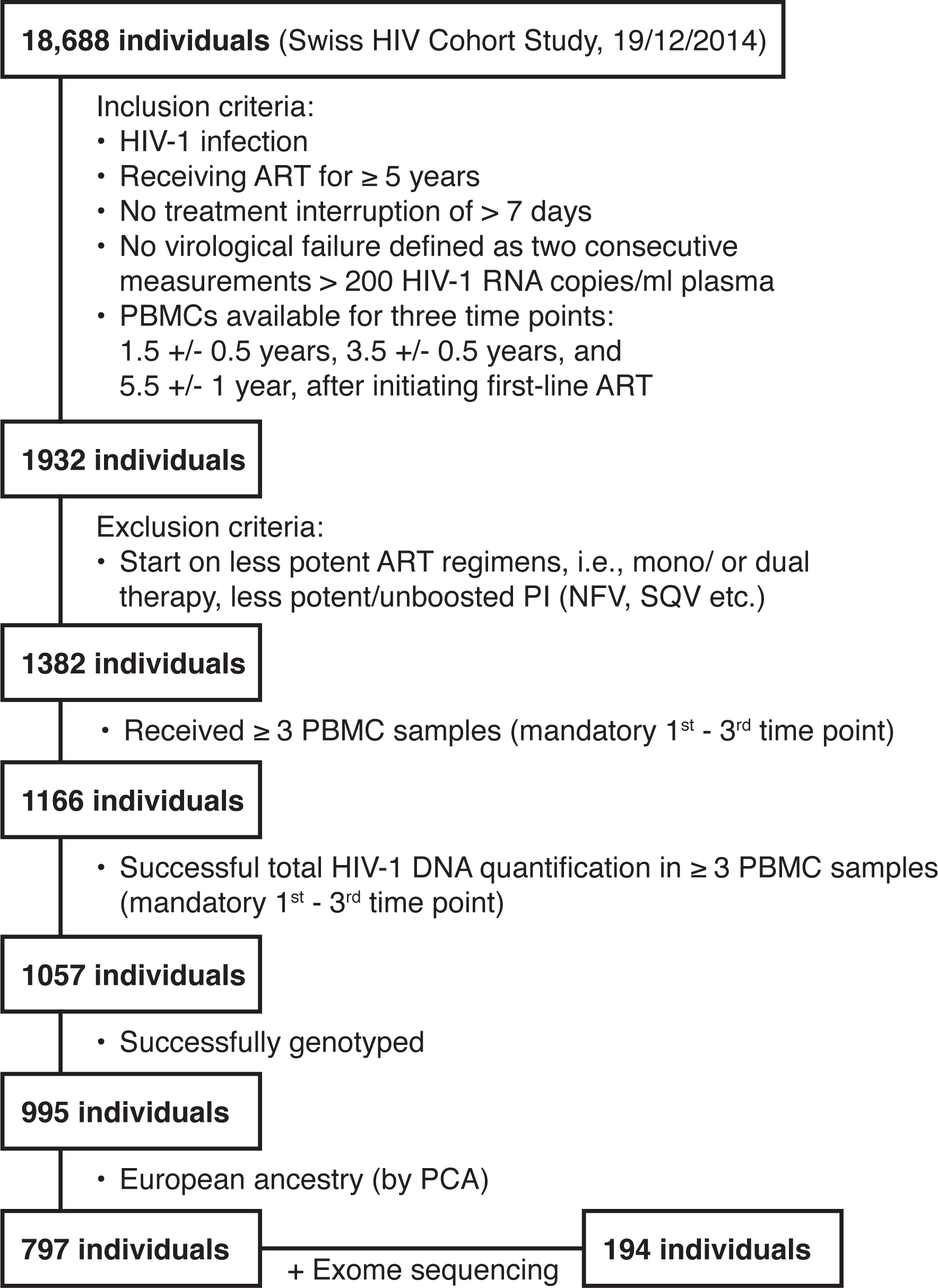
Patient selection flowchart. Specific inclusion and exclusion criteria are listed for each selection step. ART (antiretroviral therapy); PBMCs (peripheral blood mononuclear cells); PI (protease inhibitor); PCA (principal component analysis).

### Quantification of total HIV-1 DNA

The collection of longitudinal cryopreserved peripheral blood mononuclear cells (PBMCs) from eligible participants and the quantification of total HIV-1 DNA by droplet digital PCR has been described previously along with the calculation of the reservoir decay rate [28]. Briefly, this study utilized total HIV-1 DNA quantifications from three time points at a median of ∼1.5 years, ∼3.5 years, and ∼5.4 years after initiation of ART.

### Genotyping and genome-wide association analyses

Genome-wide genotyping data were obtained from previous GWAS that used various microarrays, including the HumanCore-12, HumanHap550, Human610, Human1M and Infinium CoreExome-24 BeadChips (Illumina Inc., San Diego, CA, USA), or generated from DNA extracted from peripheral blood mononuclear cells using the HumanOmniExpress-24 BeadChip (Illumina Inc., San Diego, CA, USA).

Genotypes from each genotyping array were filtered and imputed separately, with variants first flipped to the correct strand with BCFTOOLS (v1.8) according to the human GRCh37 reference genome and filtered based on a less than 20% deviation from the 1000 genomes phase 3 EUR reference panel. Genotypes were phased, and missing genotypes were imputed with EAGLE2 [41] and PBWT [42] respectively, using the 1000 Genomes Project Phase 3 reference panel on the Sanger Imputation Service [43]. Study participants were filtered based on European ancestry as determined by principal component analysis (PCA) using EIGENSTRAT (v6.1.4) [44] and the HapMap project [45] as reference populations (Figure S1A). Imputed variants were filtered by minor allele frequency (MAF) < 5%, missingness > 10%, deviation from Hardy-Weinberg equilibrium (P_HWE_ < 1e-6) and imputation quality score (INFO < 0.8). The remaining genotypes were then combined using PLINK (v1.90b5) [46] prior to analyses.

To carry out the GWASs, genome-wide genotypes were tested for association with each of the two study phenotypes (reservoir size or reservoir decay rate) in two separate genome-wide association analyses. Statistical significance was set to the standard genome-wide significance threshold of P < 5e-8 to correct for multiple testing. The associations were computed using linear mixed models with genetic relationship matrixes calculated between pairs of individuals according to the leave-one-chromosome-out method as implemented in GCTA mlma-loco (v1.91.4beta) [47,48], only including age and sex as covariates, to avoid masking of true associations by confounders. To further assess the contribution of variables previously shown to be associated with either reservoir size or decay rate, we ran multiple genome-wide association analyses, each including age, sex, and one single covariate, for each of the two study phenotypes. Finally, we conducted a GWAS including all the covariates except those showing mutual correlations. These covariates included time on ART, time to viral suppression, infection stage (acute or chronic), HIV-1 RNA pre-ART, last CD4+ T cell count pre-ART, HIV-1 subtype, transmission group, and occurrence of blips or low-level viremia during treatment.

Classical HLA alleles at the four-digit level and variable amino acids within HLA proteins were imputed using SNP2HLA (v1.03) with the T1DGC reference panel consisting of 5,225 individuals of European ancestry [49]. Association analyses with the imputed HLA alleles and multi-allelic amino acids was performed using linear regressions in PLINK and multivariate omnibus tests, respectively. For all HLA analyses age, sex and the first principal component was included as covariates.

Genotypes at specific loci, i.e. the CCR5Δ32 deletion (rs333) and the HLA-B*57:01 allele, known to influence the setpoint viral load (spVL) [50,51], available from genome-wide genotyping data, were tested for association with the reservoir size and its decay rate in 797 patients. High quality genotyping information on the CCR5Δ32 deletion was available for most individuals (N = 687), while all had available HLA information.

### Exome sequencing and analysis

All coding exons were captured using either the Illumina Truseq 65 Mb enrichment kit (Illumina Inc., San Diego, CA, USA) or the IDT xGen Exome Research Panel v1.0 (Integrated DNA Technologies Inc., Coralville, IA, USA) and sequenced on the Illumina HiSeq4000. Sequence reads were aligned to the human reference genome (GRCh37) including decoys with BWA-MEM (v0.7.10) [52]. PCR duplicates were flagged using Picard tools (v2.18.14) and variant calling performed using GATK (v3.7) [53].

To ensure a high-quality variant set across capture kit batches, all samples were merged and variants filtered based on sequencing depth (DP ≥ 20) and genotype quality (GQ ≥ 30) using BCFTOOLS (v1.8). Furthermore, individual genotypes were set as missing in cases of low depth (DP < 10) or low quality (GQ < 20). The effect of the included variants was annotated with SnpEff (v4.3T) [54].

For single variant association analysis, the VCF file was converted to PLINK format using BCFTOOLS and PLINK. Only variants with a MAF above 5%, missingness per variant below 5% and absence of severe deviation from Hardy-Weinberg equilibrium (P_HWE_ > 1e-6) were retained for the subsequent association analyses using PLINK. Sex, age and the first principal component were included as covariates. Only individuals of European descent were retained for the analyses, as determined by PCA (Figure S1B).

The combined effect of rare protein-altering variants (MAF < 5%), defined as either missense, stop-gain, frameshift, essential splice variant or an indel by SnpEff, on the reservoir size and decay rate was analyzed using optimal sequence kernel association tests (SKAT-O) [55]. For the decay rate, individuals were split into two groups due to the non-normal distribution; one exhibiting a very high decay over time (< −0.03 −log10(DNA)) and another with a stable reservoir size (≥ −0.03 and ≤ 0.03 −log10(DNA)). For this case-control analysis we used the SKATbinary function with linear weighted variants as implemented in the SKAT R package. In both cases, the analyses were adjusted for age, sex, and the first principal component.

Classical HLA class I and II alleles at the four-digit level were imputed from the exome sequencing data using HLA*LA [56]. All reads mapping to the MHC region or marked as unmapped were extracted using Samtools (v1.8) and used as input into HLA*LA. For association analyses, the 4-digit HLA alleles were extracted and analyzed using PyHLA [57] assuming an additive model, a minimum frequency of 5% and including age, sex and the first principal component as covariates.

### Copy number variation

Copy number variations (CNVs) were called from exome sequencing data using CLAMMS [58]. CNVs were called for all samples in batches according to the exome capture kit used. Within batches, samples were normalized based on coverage and potential intra-batch effects adjusted for through the use of recommended mapping metrics extracted with Picard tools (v2.18.14). After CNV calling, samples with the number of CNVs two times above the median were excluded (N = 2). CNV association analyses were performed for duplications and deletions separately for common CNVs (frequency > 5 %) with PLINK adjusting for age and sex. Potential rare CNVs (frequency < 5%) impacting immune related genes were examined by overlapping called CNVs with curated immune-related genes from Immport [59] which were also listed as protein coding in GENCODE (v25).

### Statistical analyses

All statistical analyses were performed using the R statistical software (v3.5.2), unless otherwise specified.

## RESULTS

### Host genetic determinants of the reservoir size and long-term dynamics

To investigate the effects of host genetic variation on the size of the HIV-1 reservoir 1.5 years after ART initiation and its long-term dynamics under ART over a median duration of 5.4 years, we performed a GWAS, including 797 well-characterized HIV-1 positive individuals. All study participants were enrolled in the SHCS and were of European ancestry with longitudinal total HIV-1 DNA measurements available (Table 1). The median HIV-1 reservoir size was 2.76 (IQR: 2.48-3.03) log10 total HIV-1 DNA copies/1 million genomic equivalents measured ∼1.5 years after initiation of ART (Figure S2A). The median decay rate between 1.5-5.4 years after initiation of ART was −0.06 (IQR: −0.12-0.00) log10 total HIV-1 DNA copies/1 million genomic equivalents per year (Figure S2B). With our sample size we had 80% power to detect variants with a MAF of 10% explaining at least 5% of the variance in HIV-1 reservoir size or decay rate [60].

**Table 1.** Patient characteristics.

|  |  |
| --- | --- |
| **Total number of individuals** |  |
| Genotyped | 797 |
| Genotyped + exome sequenced | 194 |
| **Age at first HIV-1 DNA sample in years** |  |
| median (IQR) | 44 (38, 50) |
| **Sex** |  |
| Female | 123 (15.4%) |
| Male | 674 (84.6%) |
| **Transmission group** |  |
| HET | 241 (30.2%) |
| IDU | 77 (9.7%) |
| MSM | 448 (56.2%) |
| Other | 31 (3.9%) |
| **HIV-1 subtype** |  |
| B | 550 (69.0%) |
| Non-B | 128 (16.1%) |
| Unknown | 119 (14.9%) |
| **Occurrence of blips or low-level viremia** |  |
| Blips | 200 (25.1%) |
| Low-level viremia | 68 (8.5%) |
| None | 529 (66.4%) |
| **Time on ART** |  |
| median (IQR) | 1.50 (1.28, 1.69) |
| **Infection stage** |  |
| Acute | 140 (17.6%) |
| Chronic | 657 (82.4%) |
| **Time to viral suppression** |  |
| median (IQR) | 0.34 (0.23, 0.51) |
| **Log<sub>10</sub> HIV-1 plasma RNA pre-ART per mL** |  |
| median (IQR) | 480 (248, 684) |
| **CD4+ cell count pre-ART cells/μL blood** |  |
| median (IQR) | 186 (90, 270) |
| **HIV-1 reservoir size** |  |
| median (IQR) | 2.76 (2.48, 3.03) |
| **HIV-1 reservoir decay rate** |  |
| median (IQR) | -0.06 (-0.12, -0.00) |

Transmission group indicates the self-reported route of infection (heterosexual (HET), intravenous drug usage (IDU), men who have sex with men (MSM), and other (including transfusions and unknown)). The occurrence of viral blips was defined by measurements of ≥
50 HIV-1 RNA copies/mL plasma within a 30-day window. Individuals with consecutive measurements of $\geq 50$ HIV-1 RNA copies/mL plasma for longer durations were classified as exhibiting low-level viremia. Time to viral suppression was the time from initiation of ART to the first viral load measurement below 50 copies/mL HIV-1 plasma RNA. HIV-1 reservoir size was measured in $\log_{10}$ total HIV-1 DNA/1 million genomic equivalents $\sim 1.5$ years after initiating ART. The HIV-1 reservoir decay rate was based on the three measurements of total HIV-1 DNA levels taken at the median of 1.5, 3.5 and 5.4 years after initiation of ART.

First, we performed GWAS using age and sex as covariates. We did not observe any genome-wide significant variant (P < 5e-8) associated with either HIV-1 reservoir size or long-term dynamics (Figure 2, S3). However, as we have previously determined, multiple factors are associated with the HIV-1 reservoir size and its decay rate [28], some of which are correlated (Figure S4). Thus, we performed additional analyses iteratively including these factors to test whether they could mask genetic associations. We ran multiple GWAS each adjusting for age, sex, plus one of the associated covariates, as well as all of the covariates together. The addition of the covariates did not have any significant effect on the results nor the genome-wide inflation factor (lambda) (Table S1).

**Figure 2.**
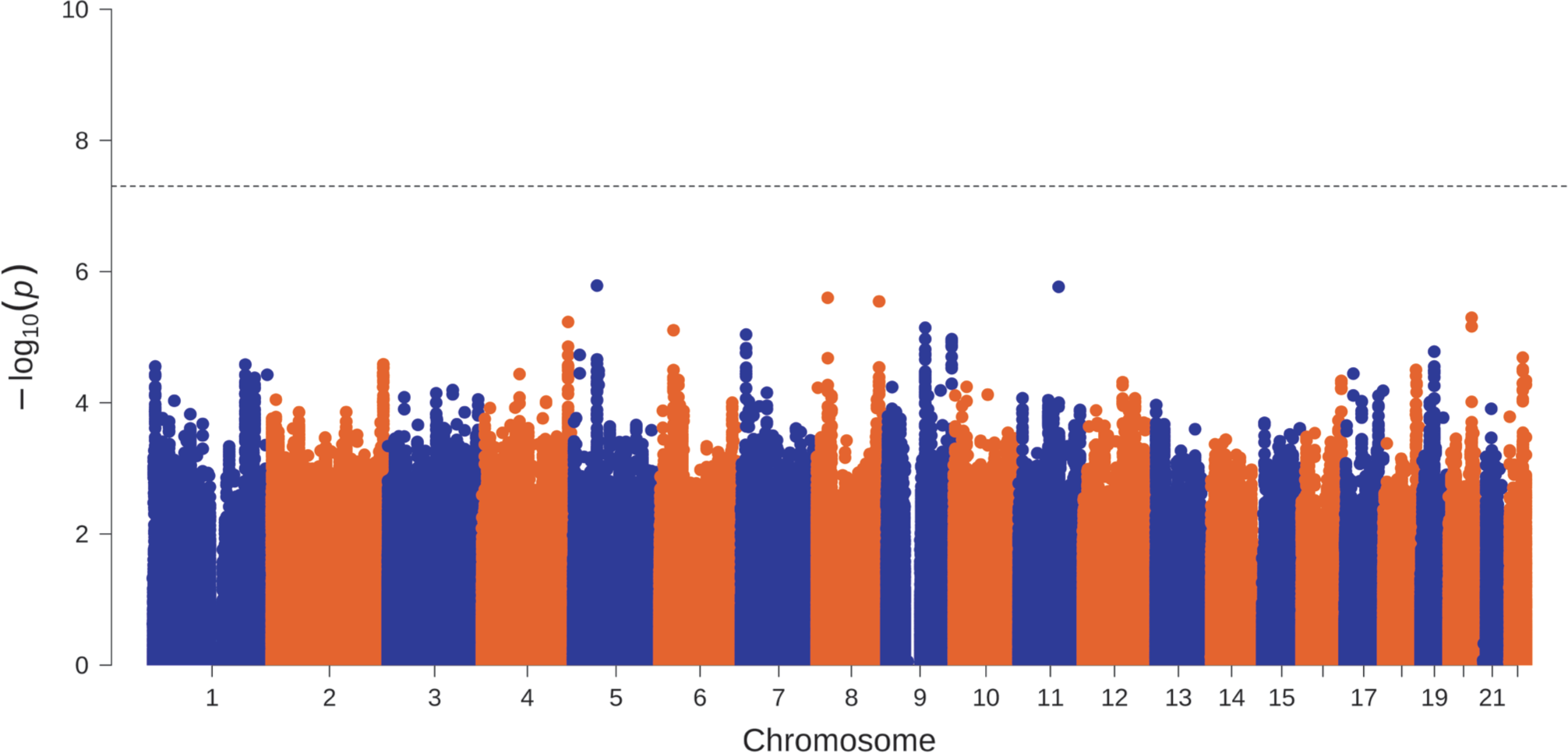
Association results with HIV-1 reservoir size. Manhattan plot with association p-values (-log_10_(P)) per genetic variant plotted by genomic position. Dashed line indicates the threshold for genome-wide significance (P = 5e-8). No variants were found to be genome-wide significant.

Genetic variation in the HLA region has previously been associated with multiple HIV-related outcomes, including spVL [51]. To test whether specific HLA variants were associated with reservoir size or long-term dynamics, we imputed the HLA alleles and amino acids for all 797 individuals from the genotyping data. In line with the previous results, we did not observe any genome-wide significant associations with any HLA allele or amino acid.

### Impact of protein-coding and rare variants

To assess the impact of rare variants as well as protein-coding variants missed by genotyping arrays on the HIV-1 reservoir size and long-term dynamics, we performed exome sequencing in 194 of the 797 study participants. Patients were selected at the two extremes of the observed reservoir decay rate: either very rapid, or absent (no change in reservoir size over ∼5.4 years), while individuals with increasing HIV-1 reservoir sizes were excluded (N=12). Thus, the long-term dynamics phenotype was binarized for subsequent analyses of the decay rate, while the HIV-1 reservoir size phenotype remained normally distributed (Figure S5).

To ensure that no common variants, missed by the genotype chips, were associated with the HIV-1 reservoir size or long-term dynamics, we performed a GWAS for common variants using age and sex as covariates. As with the genotyping data, we observed no genome-wide significant variants for either phenotype (Figure S6).

We then examined the potential role of rare variants (MAF < 5%) with a functional impact defined as either missense, frameshift, stop gained, splice acceptor or donor. Since HIV-1 primarily infects CD4+ T cells, we only included variants within genes expressed in these cells as determined by Gutierrez-Arcelus *et al*. [61]. The significance threshold after correcting for the number of tests performed was P = 1.21e-5. We did not observe any significant associations for either the HIV-1 reservoir or the decay rate. The *AMBRA1* gene showed the strongest association with HIV-1 reservoir size (P = 4.15e-5, not significant) (Figure S7).

To confirm the lack of HLA association seen with the genotyping data, we imputed the HLA haplotypes from the exome data using HLA*LA. Again, we did not observe any significant HLA association with the study outcomes.

### Copy number variations

To examine the role of large exonic CNVs not captured by standard genotyping and exome pipelines, we called CNVs from the mapped sequencing reads of the exome samples using the software CLAMMS. The contribution of common CNVs to HIV-1 reservoir size and long-term dynamics was analyzed by association analyses including age, sex and the first principal component as covariates. No significant association was observed after Bonferroni correction (Figure S8). We also searched for rare CNVs in curated immune-related genes from Immport [59] but did not discover any suggestive immune-related CNVs.

### Influence of HLA-B*57:01 and the CCR5Δ32 deletion on reservoir size and long-term dynamics

We have previously shown that pre-ART RNA viral load levels are associated with the HIV-1 reservoir size and the occurrence of blips [28]. The HLA-B*57:01 allele and the CCR5Δ32 deletion are well known genetic variants influencing HIV-1 spVL [50,51], and could thus also be associated with the with the HIV-1 reservoir size or its decay rate. However, we did not observe any nominal association (all P > 0.05) with either reservoir size or its long-term dynamics for HLA-B*57:01 and CCR5Δ32 (Figure S9).

## DISCUSSION

We used a combination of genomic technologies to assess the potential role of human genetic factors in determining both the HIV-1 reservoir size and its long-term dynamics in a well-characterized, population-based cohort. We studied 797 HIV-1-positive individuals of European origin under suppressive ART over a median of 5.4 years, for whom extensive clinical data are available, allowing detailed characterization and correction for potential confounders [28]. We measured the HIV-1 reservoir size at three time points and selected two phenotypes for our genomic study: the reservoir size at ∼1.5 years after ART initiation and the slope of the reservoir decay rate calculated over the three time points. Previous HIV host genetic studies mostly focused on phenotypes reflecting the natural history of HIV-1 infection, prior to ART initiation, including spontaneous viral control and disease progression [30–37]. A single study specifically tested for associations between common genetic variants and the amount of intracellular HIV-1 DNA, measured at a single time point during the chronic phase of infection prior to initiation of any antiretroviral therapy [38]. Here, in contrast, we longitudinally assessed samples collected from patients under suppressive ART to search for human genetic determinants of the long-term dynamics of the HIV-1 reservoir during treatment.

We first conducted a GWAS on 797 individuals to test for association between common genetic variants and the phenotypes. Given the small proportion of non-European subjects in the initial study cohort, we only included patients of European ancestry to avoid any false positive associations or masking of true positive associations due to different allele frequencies in small proportions of individuals belonging to different subpopulations (Figure 1) [62]. Regardless of including or not independent covariates other than the standard ones (i.e., sex and age), no genetic variant reached the genome-wide significance threshold for association with any of the two phenotypes. This may reflect a small effect size of genetic variants on the HIV-1 reservoir size and decay rate. We acknowledge that a larger sample size and thus increased statistical power may allow detecting genetic variants with a smaller effect size associated with the phenotypes. However, it should be noted that this study is by far the largest today that has investigated the size and decay of the HIV-reservoir in well characterized and well suppressed HIV-positive individuals over a longer time period. Alternatively, the control of the HIV-1 reservoir size and its long-term dynamics may be under the control of viral or host factors other than the individual germline genetic background. A previous report from our group had shown a correlation between viral blips during the first 1.5 years of suppressive ART and the HIV-1 reservoir size 1.5 years after ART initiation, and between viral blips after 1.5-5.4 years of suppressive ART or low-level viremia and a slower decay rate [28]. Importantly, viral blips are generally thought to reflect transient increases in viral replication, and probably occur under multifactorial influence from viral and host factors [63–70], with these latter possibly including, but not being limited to, germline genetic variation. The biological relations between viral reservoir, decay rate, viral blips, and the contribution of the individual genetic background still need full elucidation.

Standard GWAS is designed to detect associations with common genetic variants (i.e., with a MAF of at least 0.05), with little power to investigate the role of rare variants. Thus, to further assess the contribution of rare variants in individuals at the extreme of the decay rate distribution, we used exome sequencing in a selected subset of 194 study participants with very high decay rate, or conversely, a stable reservoir size over time (Figure S5). Here again, our analyses did not detect significant associations with the phenotypes. Although not reaching statistical significance, a rare genetic variant with potential functional impact in *AMBRA1* had a p-value for association just below the corrected threshold. The expression of *AMBRA1*, a core component of the autophagy machinery, has previously been associated with long-term viral control in HIV-1 non-progressors [71]. Future studies may further elucidate whether genetic variation in *AMBRA1* may account for inter-individual differences in the long-term dynamics of the HIV-1 reservoir.

Large deletions or duplications of genomic material may be implicated in human phenotypes, with CNVs impacting the exonic regions being more likely to have a functional role. Thus, we further investigated whether any common or rare CNV spanning exonic regions was associated with the phenotype. Again, no CNV was statistically associated with the phenotypes both in the exome-wide analyses and in analyses focused on immune-related genes.

An inherent limitation of our exome-based association analyses was their inability to detect rare variants outside the coding or splice-site regions. The exonic regions account for approximately 1-2% of the whole human genome. Because many regulatory sequences are located in extra-genic sites, our analysis did not fully investigate the role of highly conserved, non-coding genetic regions in influencing the phenotypes linked to HIV-1 latency.

Additionally, we focused on specific genetic variants, i.e., the HLA haplotypes and the CCR5Δ32 deletion, previously demonstrated to have a role in HIV-1 related phenotypes [30,51]. Indeed, previous studies unraveled a robust association between variation in the HLA region and the HIV-1 spVL [30]. Likewise, heterozygosity for the CCR5Δ32 deletion has been shown to influence spontaneous HIV-1 control [51]. Thus, we imputed HLA genotypes from genotyping and exome data, and studied the CCR5Δ32 deletion, without, however, detecting any significant associations with the phenotypes or the covariates (Figure S9). Specifically, we found no correlation between HLA genotypes and HIV-1 RNA plasma levels prior to ART initiation, apparently contrasting with the previous findings of an association between HLA-B*57:01 haplotype and spVL. This probably reflects historical changes in the therapeutic approach following a diagnosis of HIV-1 infection, given that ART is currently initiated soon after clinical diagnosis, before most patients reach a stable plateau of plasma viral load.

In our study, the quantification of the reservoir size at different time points may have been influenced by factors as, for example, blips and low-level viremia, which may have reduced our ability to detect significant genetic effects. It is also possible that, in the future, novel methods to assess the viral reservoir will allow the detection of significant contributions of genetic factors [72]. So far, it remains unanswered whether the initial response to acute infection, the containment of ongoing replication, and the control of latently infected cells are under the influence of the same or different molecular networks. It needs to be noted that in previous work we have shown that host genetic factors as defined by GWAS did not explain the severity of symptoms during acute HIV-infection, although severity of symptoms correlated well with viral load and CD4 cell counts [73].

In conclusion, our study suggests that human individual germline genetic variation has little, if any, influence on the control of the HIV-1 viral reservoir size and its long-term dynamics. Complex, likely multifactorial biological processes govern HIV-1 viral persistence. Larger studies will possibly clarify the role of common or rare genetic variants explaining small proportions of the variability of the phenotypes related to viral latency.

## Data Availability

The datasets generated during and/or analyzed during the current study are not publicly available due to privacy reasons, the sensitivities associated with HIV infections, and the representativeness of the dataset, but is available on request.

## Author contributions

N.B., J.B., V.R., R.D.K., H.F.G., K.J.M., and J.F. contributed to the conception and design of the study. C.v.S., V.V., K.N., Y.I.K., and K.J.M. contributed to the acquisition of data. A.B., C.W.T., N.B., T.T., S.P., M.W., R.D.K., and J.F. contributed to the analysis and interpretation of data. J.B., M.P., T.K., S.Y., M.B., A.R., P.S., E.B., M.C., H.F.G., and the members of the Swiss HIV Cohort Study (SHCS) conceived and managed the cohort, collected and contributed patient samples and clinical data. A.B., C.W.T., and J.F. contributed to the drafting the article. All authors read and approved the final manuscript.

## Acknowledgements

We thank the patients for participating in the SHCS, the study nurses and physicians for excellent patient care, A. Scherrer, A. Traytel, and S. Wild for excellent data management and D. Perraudin and M. Amstutz for administrative assistance.

This work was funded within the framework of the Swiss HIV Cohort Study (SNF grant# 33CS30_177499 to H.F.G.). The data were gathered by the Five Swiss University Hospitals, two Cantonal Hospitals, 15 affiliated hospitals and 36 private physicians (listed in http://www.shcs.ch/180-health-care-providers). The work was furthermore supported by the Systems.X grant # 51MRP0_158328 (to N.B., J.B., J.F.,V.R., R.D.K., M.W., H.F.G. and K.J.M.), by SNF grant 324730B_179571 (to H.F.G.), SNF grant SNF 310030_141067/1 (to H.F.G. and K.J.M.), SNF grants no. PZ00P3-142411 and BSSGI0_155851 to R.D.K., the Yvonne-Jacob Foundation (to H.F.G.), the University of Zurich’s Clinical Research Priority Program viral infectious disease, ZPHI (to H.F.G) and the Vontobel Foundation (to H.F.G. and K.J.M.). M.W. is partially supported by the NCCR MARVEL, funded by the Swiss National Science Foundation.

Members of the Swiss HIV Cohort Study:

Anagnostopoulos A, Battegay M, Bernasconi E, Böni J, Braun DL, Bucher HC, Calmy A, Cavassini M, Ciuffi A, Dollenmaier G, Egger M, Elzi L, Fehr J, Fellay J, Furrer H, Fux CA, Günthard HF (President of the SHCS), Haerry D (deputy of “Positive Council”), Hasse B, Hirsch HH, Hoffmann M, Hösli I, Huber M, Kahlert CR (Chairman of the Mother & Child Substudy), Kaiser L, Keiser O, Klimkait T, Kouyos RD, Kovari H, Ledergerber B, Martinetti G, Martinez de Tejada B, Marzolini C, Metzner KJ, Müller N, Nicca D, Paioni P, Pantaleo G, Perreau M, Rauch A (Chairman of the Scientific Board), Rudin C, Scherrer AU (Head of Data Centre), Schmid P, Speck R, Stöckle M (Chairman of the Clinical and Laboratory Committee), Tarr P, Trkola A, Vernazza P, Wandeler G, Weber R, Yerly S.

